# In-hospital stay of anemic patients (70-90 g.L^-1^) in the ED with/without transfusion: a single-center propensity-matched study

**DOI:** 10.1101/2024.09.11.24313465

**Authors:** Fabien Coisy, Clémence Anselme, Radjiv Goulabchand, Laura Grau-Mercier, Thibaut Markarian, Xavier Bobbia, Romain Genre-Grandpierre

**Affiliations:** UR UM 103 (IMAGINE), Department of Emergency Medicine, CHU Nîmes, University of Montpellier, Nîmes, France; Department of Emergency Medicine, CHU Nimes, University of Montpellier, Nîmes, France; IDESP, INSERM, Department of Internal Medicine, CHU Nîmes, University of Montpellier, Nîmes, France; Department of Emergency Medicine, Timone University Hospital, Marseille, France; UR UM 103 (IMAGINE), Department of Emergency Medicine, Montpellier University Hospital, Montpellier University, Montpellier, France

**Keywords:** Blood transfusion, Length of stay, Anemia, Emergency Department, Hospital

## Abstract

**Background and importance:** A quarter of patients presenting to the emergency department (ED) have anemia. Although red blood cell (RBC) transfusion is routinely used in symptomatic anemia, there is no evidence on the benefit of blood transfusion in hemodynamically stable patients in the ED for patients requiring hospitalization.

**Objective:** The study aimed to compare in-hospital length of stay (LOS) of patients with anemia between 70 and 90 g.L-^1^ transfused or not in ED.

**Design:** Retrospective single-center study

**Settings and participants:** All adult patients admitted to the ED of our university hospital with an initial hemoglobin level between 70 and 90 g.L^-1^, without hemorrhagic shock, who were hospitalized after ED admission.

**Outcome measures and analysis:** A propensity score, comprising hemoglobin level, Charlson’s comorbidity index, clinical signs of anemia, the chronicity of anemia and hospitalization department was used to compare the LOS of patients transfused versus non-transfused in the ED.

**Intervention:** RBC transfusion in the ED

**Main results:** From January 1st to December 31st, 2022, 1 169 patients were screened of whom 569 (49%) were excluded, mostly due to discharge without hospitalization. The remaining 564 (48%) patients had a median age of 77 [68; 85] and 240 (43%) were women. Finally, 127 (23%) patients were transfused in ED. Transfused patients received more units of RBC during the whole hospitalization period (4 [3; 5] versus 2 [1; 3] than non-transfused patients (p< 0.01)). After propensity score matching, median LOS was 9 [5; 19] days for ED transfused patients and 8 [5; 15] days for non-ED transfused patients (median difference= -1 95% CI [-3; 2]; p=0.45).

**Conclusion:** In patients with non-life-threatening anemia, RBC transfusion in the ED does not appear to reduce in-hospital LOS compared with transfusion in inpatient departments. Further studies are needed to identify patients requiring transfusion in ED.

## Introduction

Anemia, defined by World Health Organization as a hemoglobin level under 120 g.L^-1^ for women and 130 g.L^-1^ for men, is a worldwide concern [1]. Anemia is a public health concern as it worsens the prognosis of several diseases such as heart-failure and may increase in-hospital length of stay (LOS) [3]. Symptoms of anemia are unspecific, and many patients may be asymptomatic [4]. The quickest therapeutic response to symptomatic anemia is packed red blood cell (RBC) transfusion. Current recommendations advocate a restrictive transfusion strategy, meaning not transfusing over a hemoglobin threshold of 70 g.L^-1^ in hemodynamically stable patients [5,6]. However, it remains uncertainty concerning the transfusion of patients having hemoglobin level between 70 and 90 g.L^-1^.

In emergency departments (ED), 12 to 25% of admitted patients may present with anemia [7,8]. The proportion of patients with symptomatic anemia remains unclear, as patients in the ED often present complex diseases with overlapping symptoms such as fatigue, dyspnea, or tachycardia [5]. Conflicting reports suggest over-transfusion strategies in ED [9], increasing the risk of wrong blood in tube errors, and transfusion related side effect [10,11]. Nevertheless, blood transfusion in ED could account for 10 to 28% of in-hospital transfusions, annually [12,13]. Studies on RBC transfusion appropriateness shows that it is ranged between 20 and 40% in ED [13, 14].

No study in high-income country showed a benefit for patient with anemia and without life-threatening condition, to be transfused in ED. We hypothesize that transfusing hemodynamically stable patients requiring hospitalization, admitted to the ED with anemia between 70 and 90 g.L^-1^ would alter in-hospital LOS. We aimed to determine whether there was a difference in in-hospital LOS between patients transfused in the ED and those who were not.

## Methods

### Study settings

This study was a retrospective single-center study in the university hospital of Nîmes, France conducted between January 1^st^ to December 31^st^, 2022. This ED received 119 301 patients in 2022. The local ethics committee of Nimes’ university hospital approved the study (IRB 23.05.02). A non-opposition letter was sent to patients or their relative if patients were dead during hospitalization, explaining the aims of the study and the possibility to oppose to data collection. Data were collected from May 30^th^, 2023, to September 30^tht^ 2023. Authors had access to information that could identify individual participants during data collection, that were not collected.

### Objectives

The primary outcome was the difference in LOS, measured in days as the time between ED admission and hospital discharge date, between patients presenting in ED with anemia between 70 and 90 g.L^-1^ with or without transfusion with RBC in the ED. Secondary outcomes were the in-hospital mortality between the two groups, the difference in long hospitalization (in-hospital LOS above the cohort median), and differences of LOS considering the number of RBC transfused in the ED, in ward and in total.

### Population

The study population involved ≥18-year-old patients who presented to the ED with a biological diagnosis of anemia, characterized by hemoglobin levels between 70 and 90 g.L^-1^, who were subsequently hospitalized, whatever the reason of hospitalization was. The patients were clinically stable. Patients with hemorrhagic shock, defined as acute blood loss, either external or internal, associated with hemodynamic failure and hyperlactatemia greater than 2 mmol.L^-1^ of traumatic or non-traumatic origin, were not included. Patients requiring surgical intervention, radiological or endoscopic procedures, or emergency hemostatic procedures, as well as pregnant women, were excluded. Patients returning home after ED admission were excluded. Finally, for patients presenting multiple times in ED with anemia, only the first admission was analyzed.

### Data collection

Data were collected retrospectively from patients’ electronic files: age, sex, reason for consultation, arterial blood pressure, heart rate, oxygen saturation, and Charlson’s comorbidity index. Symptoms of anemia were defined by the presence of asthenia, dyspnea, chest pain, dizziness, syncope or blood loss within the ED admission causes. First ED biological results were collected: hemoglobin level, mean corpuscular volume, iron and transferrin saturation coefficient, platelet count, leukocyte count and serum creatinine level. The presence of RBC transfusion and the number of RBC units transfused in ED were also collected. The total ED LOS was collected and defined as the time between ED admission and ED discharge. Boarding, defined by “holding admitted patients in the ED, often in hallways, while awaiting an inpatient bed” was recorded as an informatic transfer in the hospitalization waiting zone [15]. Patients transfused in the hospitalization waiting zone were considered as transfused in ward, as if a bed was available, they would not have stayed in ED. Hospitalization ward was collected, and classed as medicine ward or intensive care unit (ICU). Transfusion-related side effects (dyspnea, tachycardia, fever, hypertension, or hypotension) were recorded on the first day following ED admission. In-hospital death was also identified from medical files.

### Statistical analysis

Missing data were imputed using a linear regression model for hemodynamic parameters if the number of missing values was less than 25% of the total sample size. For in-hospital LOS, there was no missing data. Qualitative variables were expressed in absolute value and percentage. Quantitative variables were expressed in median with the first and third quartiles. Qualitative variables were compared using the Chi-square test or Fisher’s exact test depending on sample size. Quantitative variables were compared using the Mann-Whitney U test. As in-hospital death was considered as an adverse effect, adjusted hospital LOS was created, imputing maximal cohort LOS to in-hospital dead patients.

Propensity score analysis was conducted, matching according to ED hemoglobin levels, Charlson comorbidity index, clinical signs of anemia, presence of chronic anemia, and hospitalization ward. Description of propensity score is provided in supplemental material S1. Median difference and 95% confidence intervals (95%CI) were calculated for quantitative variables using resampling with a Bootstrap technique with a sample of n = 1000. Odds ratios (OR) were calculated by logistic regression for qualitative variables, along with their 95%CI. Paired groups were compared using a paired Wilcoxon test or a paired Chi-square test for qualitative variables. Comparison between more than two groups was conducted using ANOVA, followed by Tukey post-hoc test with a Holm correction for multiple testing. All tests used were two-tailed with significance set at 5%. Being an exploratory study, no number of subjects was calculated. Data were analyzed and plots were drawn using R software, version 4.2.2. (R Core Team (2022). R: A language and environment for statistical computing. R Foundation for Statistical Computing, Vienna, Austria), associated with the MatchIt, multcomp, and epiR packages.

## Results

### Population characteristics

From January 1^st^ to December 31^st^, 2022, 11 208 (9%) had anemia with less than 120 g.L^-1^. A total of 1 169 (10%) patients having anemia between 70 and 90 g.L^-1^ were screened, of which 569 (49%) were excluded: 390 (69%) patients were not hospitalized after ED admission and 55 (10%) had missing data (Figure 1). A total of 564 (48%) unique patients with anemia between 70 and 90 g.L^-1^ were analyzed. Median age of population was 77 [68; 85] years old, and 240 (43%) were women. Eleven (2%) patients had hemoglobinopathy (sickle cell anemia or thalassemia) or hydroxocobalamin malabsorption and were not transfused in ED. Median hemoglobin value at ED admission was 83 [78; 87] g.L^-1^. Main symptoms at ED presentation were fatigue, dyspnea and recent external blood loss for respectively 128 (23%), 128 (23%) and 117 (21%) patients. Median ED LOS was 13:42 [7:38; 22:52]. Median in-hospital LOS for the cohort was 10 [5; 17] days. A total of 127 (23%) patients had RBC transfusion in the ED. Among the 351 (62%) patients transfused during in-hospital stay, the median number of RBC units transfused was 2 [2; 4]. Patients baseline data are presented in Table 1.

**Figure 1:**
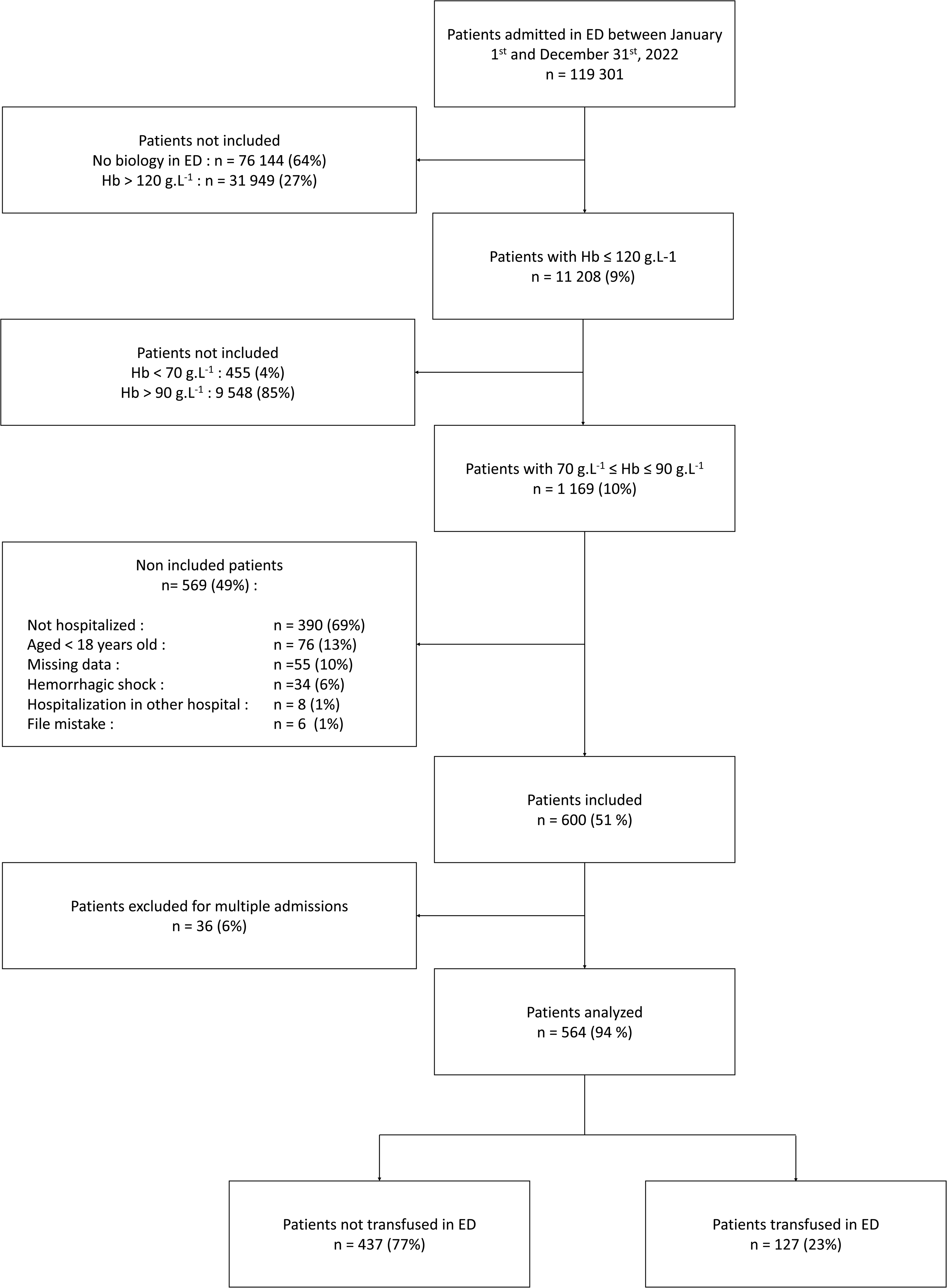
Flow chart. ED: Emergency department, Hb: hemoglobin

**Figure 2:**
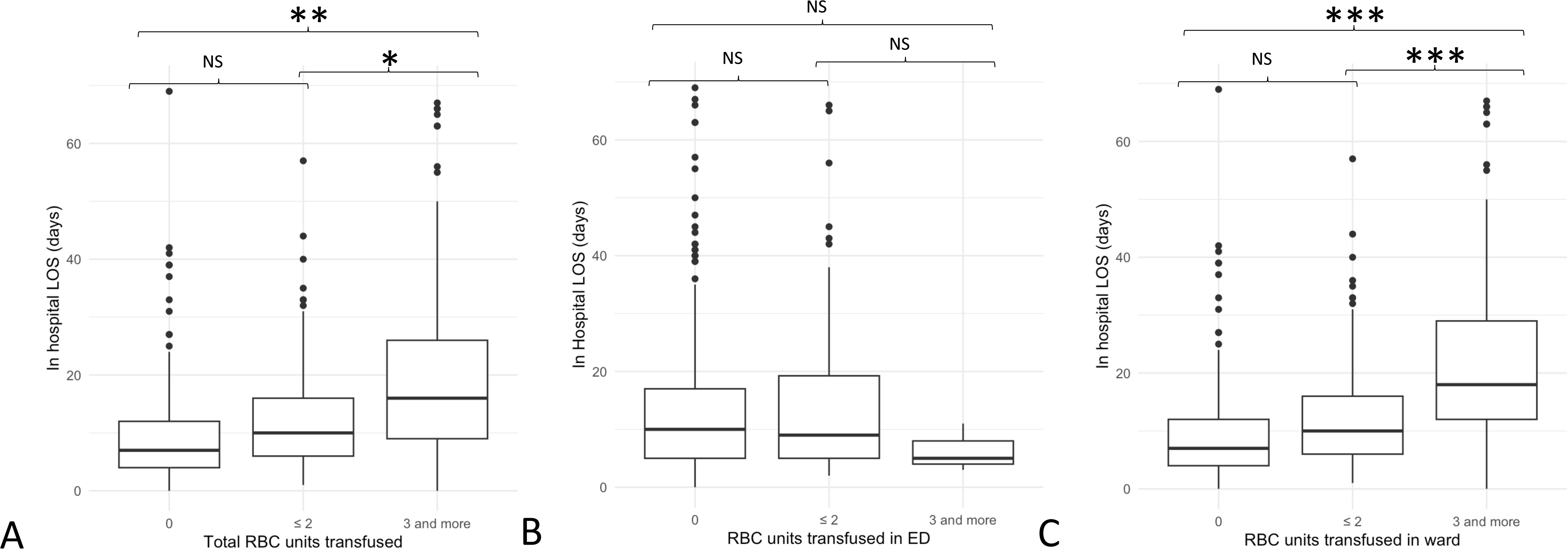
Comparison of in-hospital length of stay according to number of units and place of transfusion in the whole cohort (n = 564 patients): A. During total stay, B. In emergency department only, C. In-hospitalization only. LOS: length of stay, ED: Emergency department, RBC: red blood cell NS: not significant; * p-value < 0.05; ** p-value < 0.01; *** p-value < 0.001

**Table 1:** Characteristics of the global study population and considering the localization of their first transfusion. ED: emergency department, MCV: mean corpuscular volume, RBC: red blood cell $ Symptoms of anemia corresponded to presentation to emergency department with a complaint of asthenia or external blood loss or chest pain or syncope or dizziness or fall or fatigue or dyspnea. * Count for 351 (62%) patients: 75 (59%) transfused in ED and 276 (63%) not transfused in emergency department.

|  | Total<br>n = 564 | Non-ED<br>transfused<br>patients<br>n = 437 | ED transfused<br>patients<br>n = 127 | p-value |
| --- | --- | --- | --- | --- |
| **Demographics** |  |  |  |  |
| Age in years, m [Q1; Q3] | 77 [68; 85] | 76 [67; 86] | 78 [70; 84] | 0.95 |
| Sex, women, n(%) | 240 (43) | 189 (43) | 51 (40) | 0.54 |
| **Medical history** |  |  |  |  |
| Charlson comorbidity index, m [Q1; Q3] | 4 [2; 6] | 4 [2; 6] | 4 [2; 6] | 0.96 |
| Myocardial infarction, n(%) | 112 (20) | 74 (17) | 38 (30) | < 0.01 |
| Chronic heart failure, n(%) | 51 (9) | 37 (8) | 14 (11) | 0.38 |
| Chronic obstructive pulmonary disease, n(%) | 84 (15) | 55 (13) | 29 (23) | < 0.01 |
| Chronic kidney disease, n(%) | 211 (37) | 165 (38) | 46 (36) | 0.84 |
| Chronic anemia, n(%) | 360 (64) | 298 (68) | 62 (49) | < 0.01 |
| **Symptoms reported at ED admission** |  |  |  |  |
| Fatigue, n(%) | 128 (23) | 102 (23) | 36 (28) | 0.24 |
| Dyspnea, n(%) | 128 (23) | 99 (23) | 29 (23) | > 0.99 |
| Recent external blood loss, n(%) | 117 (21) | 55 (13) | 62 (49) | < 0.01 |
| Fall, n(%) | 60 (11) | 54 (12) | 6 (5) | 0.01 |
| Chest pain, n(%) | 25 (4) | 24 (5) | 1 (1) | < 0.05 |
| Syncope, n(%) | 14 (2) | 11 (3) | 3 (2) | > 0.99 |
| Dizziness, n(%) | 4 (1) | 2 (0) | 2 (2) | 0.22 |
| Symptoms of anemia\$, n(%) | 383 (68) | 277 (63) | 106 (83) | < 0.01 |
| **Vital parameters** |  |  |  |  |
| Systolic blood pressure in mmHg, m [Q1; Q3] | 126 [111; 137] | 126 [114; 140] | 121 [105; 132] | < 0.01 |
| Diastolic blood pressure in mmHg, m [Q1; Q3] | 66 [58; 74] | 67 [60; 75] | 62 [53; 70] | < 0.01 |
| Heart rate in beats per minute, m [Q1; Q3] | 89 [80; 100] | 89 [82; 100] | 89 [75; 99] | 0.42 |
| Oxygen saturation in %, m [Q1; Q3] | 97 [96; 98] | 97 [96; 98] | 97 [96; 98] | 0.20 |
| **Biological parameters** |  |  |  |  |
| Hemoglobin in g.L<sup>-1</sup>, m [Q1; Q3] | 83 [78; 87] | 84 [80; 88] | 77 [73; 83] | < 0.01 |
| MCV in fL, m [Q1; Q3] | 93 [86; 99] | 92 [86; 99] | 95 [88; 100] | 0.05 |
| Iron status assessed, n(%) | 76 (13) | 46 (11) | 30 (24) | < 0.01 |
| Platelets in G.L<sup>-1</sup>, m [Q1; Q3] | 233 [147;<br>340] | 226 [135;<br>344] | 254 [177;<br>330] | 0.18 |
| Leukocytes in G.L<sup>-1</sup>, m [Q1; Q3] | 9.4 [6.3; 13.5] | 9.4 [6.0; 13.6] | 9.6 [6.8; 12.8] | 0.97 |
| Serum creatinine level in $\mu\text{mol.L}^{-1}$ | 106 [73; 168] | 106 [71; 172] | 101 [78; 148] | 0.52 |
| **Hospitalization ward** |  |  |  |  |
| Medicine, n(%) | 499 (88) | 387 (89) | 112 (88) | 0.88 |
| Intensive care unit, n(%) | 65 (12) | 50 (11) | 15 (12) |  |
| **Outcomes** |  |  |  |  |
| ED LOS in hh:mm, m [Q1 ; Q3] | 13:24 [7:38 ;<br>22:52] | 12:29 [7:08 ;<br>21:50] | 17:43 [11:00 ;<br>24:40] | < 0.01 |
| Boarding, n(%) | 190 (34) | 130 (30) | 60 (47) | < 0.01 |
| More than 1 day stay in ED, n(%) | 124 (22) | 89 (21) | 35 (28) | 0.11 |
| Transfusion-related side effect, n(%) | 7 (1) | 3 (1) | 4 (3) | < 0.05 |
| Transfusion during hospital stay, n(%) | 351 (62) | 276 (63) | 75 (59) | 0.41 |
| Number of RBC units transfused in ward *, m [Q1; Q3] | 2 [1; 3] | 2 [1; 3] | 2 [1; 3] | 0.56 |
| Total number of RBC units transfused*, m [Q1; Q3] | 2 [2; 4] | 2 [1; 3] | 4 [3; 5] | < 0.01 |
| Hospital LOS in days, m [Q1; Q3] | 10 [5;17] | 10 [5; 17] | 9 [5; 19] | 0.71 |
| Adjusted hospital LOS in days, m [Q1; Q3] | 12 [6; 27] | 12 [6; 27] | 11 [6; 26] | 0.94 |
| In-hospital death, n(%) | 86 (34) | 68 (33) | 18 (38) | 0.61 |

### Primary outcome

After pairing on propensity score, 254 patients remained, 127 per group. Baseline characteristics were comparable across matched groups, and there were no missing data. The median in-hospital LOS was 8 [5; 15] for non-ED transfused patients and 9 [5; 19] days for ED transfused patients (median difference = -1; 95%CI = [-3; 2]; p = 0.45). Adjusted hospital LOS was 11 [6;26] days un non-ED transfused patients, and 11 [6; 26] days in ED transfused patients (median difference = 0; 95%CI = [-4; 4], p = 0.98). The propensity matched comparison is presented in Table 2.

**Table 2:**
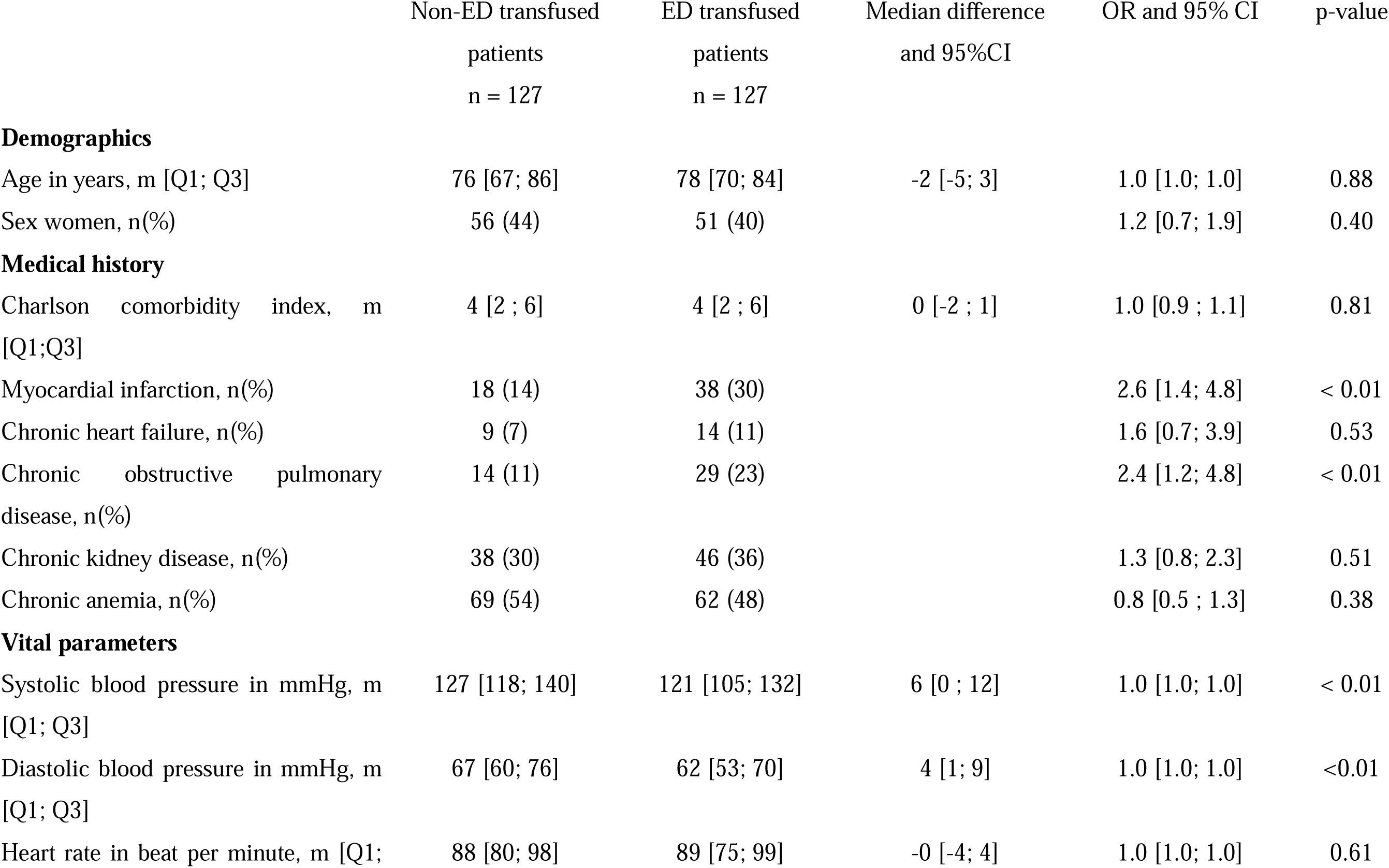
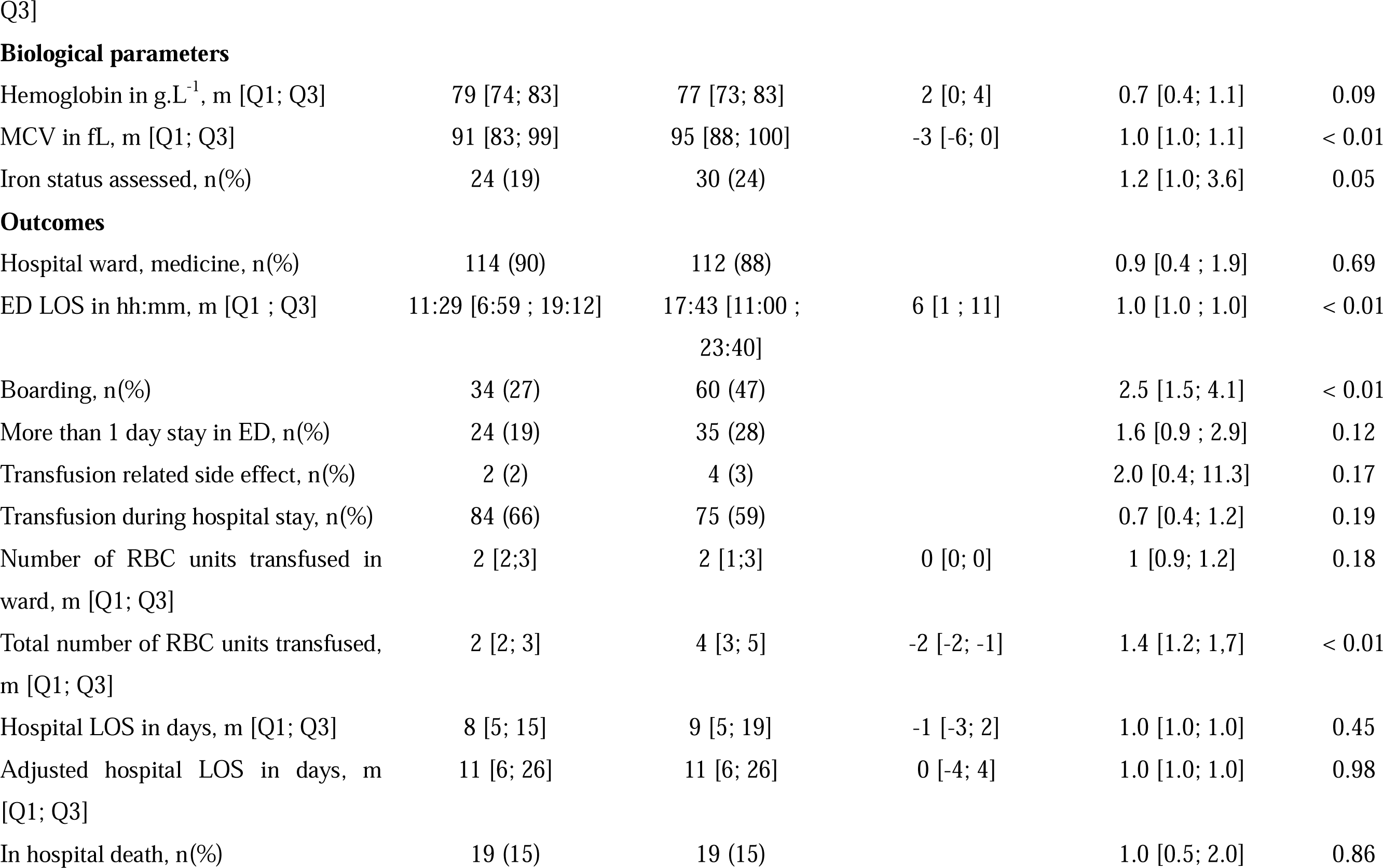
Comparison of baseline characteristics and main outcomes between patients transfused in emergency department or not, after pairing on propensity score, taking into account Charlson’s comorbidity index, hospitalization ward, symptoms of anemia, hemoglobin level and chronic anemia. OR: odds ratio, 95%CI: 95% confidence interval, ED: emergency department, MCV: mean corpuscular volume, RBC: Red blood cell

|  | Non-ED transfused<br>patients<br>n = 127 | ED transfused<br>patients<br>n = 127 | Median difference<br>and 95%CI | OR and 95% CI | p-value |
| --- | --- | --- | --- | --- | --- |
| **Demographics** |  |  |  |  |  |
| Age in years, m [Q1; Q3] | 76 [67; 86] | 78 [70; 84] | -2 [-5; 3] | 1.0 [1.0; 1.0] | 0.88 |
| Sex women, n(%) | 56 (44) | 51 (40) |  | 1.2 [0.7; 1.9] | 0.40 |
| **Medical history** |  |  |  |  |  |
| Charlson comorbidity index, m [Q1;Q3] | 4 [2 ; 6] | 4 [2 ; 6] | 0 [-2 ; 1] | 1.0 [0.9 ; 1.1] | 0.81 |
| Myocardial infarction, n(%) | 18 (14) | 38 (30) |  | 2.6 [1.4; 4.8] | < 0.01 |
| Chronic heart failure, n(%) | 9 (7) | 14 (11) |  | 1.6 [0.7; 3.9] | 0.53 |
| Chronic obstructive pulmonary disease, n(%) | 14 (11) | 29 (23) |  | 2.4 [1.2; 4.8] | < 0.01 |
| Chronic kidney disease, n(%) | 38 (30) | 46 (36) |  | 1.3 [0.8; 2.3] | 0.51 |
| Chronic anemia, n(%) | 69 (54) | 62 (48) |  | 0.8 [0.5 ; 1.3] | 0.38 |
| **Vital parameters** |  |  |  |  |  |
| Systolic blood pressure in mmHg, m [Q1; Q3] | 127 [118; 140] | 121 [105; 132] | 6 [0 ; 12] | 1.0 [1.0; 1.0] | < 0.01 |
| Diastolic blood pressure in mmHg, m [Q1; Q3] | 67 [60; 76] | 62 [53; 70] | 4 [1; 9] | 1.0 [1.0; 1.0] | <0.01 |
| Heart rate in beat per minute, m [Q1; | 88 [80; 98] | 89 [75; 99] | -0 [-4; 4] | 1.0 [1.0; 1.0] | 0.61 |
| **Biological parameters** |  |  |  |  |  |
| Hemoglobin in g.L<sup>-1</sup>, m [Q1; Q3] | 79 [74; 83] | 77 [73; 83] | 2 [0; 4] | 0.7 [0.4; 1.1] | 0.09 |
| MCV in fL, m [Q1; Q3] | 91 [83; 99] | 95 [88; 100] | -3 [-6; 0] | 1.0 [1.0; 1.1] | < 0.01 |
| Iron status assessed, n(%) | 24 (19) | 30 (24) |  | 1.2 [1.0; 3.6] | 0.05 |

Table 2: Comparison of baseline characteristics and main outcomes between patients transfused in emergency department or not, after pairing on propensity score, taking into account Charlson's comorbidity index, hospitalization ward, symptoms of anemia, hemoglobin level and chronic anemia.
|  |  |  |  |  |  |
| --- | --- | --- | --- | --- | --- |
| Hospital ward, medicine, n(%) | 114 (90) | 112 (88) |  | 0.9 [0.4 ; 1.9] | 0.69 |
| ED LOS in hh:mm, m [Q1 ; Q3] | 11:29 [6:59 ; 19:12] | 17:43 [11:00 ; 23:40] | 6 [1 ; 11] | 1.0 [1.0 ; 1.0] | < 0.01 |
| Boarding, n(%) | 34 (27) | 60 (47) |  | 2.5 [1.5; 4.1] | < 0.01 |
| More than 1 day stay in ED, n(%) | 24 (19) | 35 (28) |  | 1.6 [0.9 ; 2.9] | 0.12 |
| Transfusion related side effect, n(%) | 2 (2) | 4 (3) |  | 2.0 [0.4; 11.3] | 0.17 |
| Transfusion during hospital stay, n(%) | 84 (66) | 75 (59) |  | 0.7 [0.4; 1.2] | 0.19 |
| Number of RBC units transfused in ward, m [Q1; Q3] | 2 [2;3] | 2 [1;3] | 0 [0; 0] | 1 [0.9; 1.2] | 0.18 |
| Total number of RBC units transfused, m [Q1; Q3] | 2 [2; 3] | 4 [3; 5] | -2 [-2; -1] | 1.4 [1.2; 1.7] | < 0.01 |
| Hospital LOS in days, m [Q1; Q3] | 8 [5; 15] | 9 [5; 19] | -1 [-3; 2] | 1.0 [1.0; 1.0] | 0.45 |
| Adjusted hospital LOS in days, m [Q1; Q3] | 11 [6; 26] | 11 [6; 26] | 0 [-4; 4] | 1.0 [1.0; 1.0] | 0.98 |
| In hospital death, n(%) | 19 (15) | 19 (15) |  | 1.0 [0.5; 2.0] | 0.86 |

**Table 3:** Comparison of baseline characteristics and main outcomes between patients with or without a long-adjusted length of stay (defined by the median length of stay of the entire cohort). ED: Emergency department

| | LOS ≤ 10<br>days<br>n = 293 | LOS > 10<br>days<br>n = 271 | p-value |
| --- | --- | --- | --- |
| **Demographics** |  |  |  |
| Age in years, m [Q1; Q3] | 76 [66; 85] | 78 [70; 85] | 0.10 |
| Sex, women, n(%) | 136 (46) | 104 (38) | 0.06 |
| **Medical history** |  |  |  |
| Charlson's comorbidity index, m [Q1;Q3] | 4 [2;6] | 4 [2;6] | 0.13 |
| Chronic anemia, n(%) | 190 (65) | 170 (63) | 0.66 |
| **Biological parameters** |  |  |  |
| Hemoglobin in g.L<sup>-1</sup>, m [Q1;Q3] | 83 [78; 87] | 83 [77; 87] | 0.41 |
| Symptoms of anemia, n(%) | 200 (68) | 183 (68) | 0.86 |
| **Hospitalization ward** |  |  |  |
| Medicine, n(%) | 273 (93) | 226 (83) | < 0.01 |
| **Outcomes** |  |  |  |
| ED LOS in hh:mm, m[Q1;Q3] | 13:52 [8:07 ;<br>22:23] | 13:13 [7:22 ;<br>23:42] | 0.96 |
| Boarding, n(%) | 102 (35) | 88 (32) | 0.59 |
| ED-transfused, n(%) | 69 (24) | 58 (21) | 0.55 |
| Ward transfused, n(%) | 159 (54) | 192 (71) | < 0.01 |
| Hospital death, n(%) | 1 (0) | 85 (31) | < 0.01 |

### Secondary outcomes

In-hospital mortality was (19 (15%) in non-ED transfused patients and 19 (15%) in ED-transfused patients (OR = 1.0, 95%CI = [0.5; 2.0], p = 0.86).

Long hospitalization was defined above 10 days. Patients with and without long hospitalization had same initial hemoglobin level (83 [77; 87] vs 83 [78; 87] g.L^-1^, p =0.41) and were transfused in ED in the same proportion (58 (21%) vs 69 (24%), p = 0.96). Patients with long hospitalization were more prone to be hospitalized in ICU (45 (17%) vs 20 (7%), p < 0.01) and were more transfused in ward (192 (71%) vs 159 (54%), p < 0.01) than patients with hospitalization LOS < 10 days.

In-hospital LOS for the total population considering the number of RBC units transfused during total hospitalization was shorter for non-transfused patients than those transfused with ≥ 3 RBC units (7 [4; 12] days vs 16 [9; 26] days, p<0.01). The same trend was seen between patients transfused with 1 or 2 RBC units compared to those transfused with ≥ 3 RBC units (10 [6; 16] days and 16 [9; 26] days (p<0.05)). There was no significant difference in LOS in patients transfused with 1 or 2 units versus non-transfused (Figure 3A). In contrast, for in-hospital LOS according to the units of RBC transfused only in the ED, no difference in mean LOS was seen between any groups: 10 [5; 17] days for non-transfused patients in the ED, 9 [5; 19] days for patients transfused with 1 or 2 RBC units in the ED and 5 [4; 8] days for those transfused with ≥ 3 RBC units (ANOVA’s p-value = 0.70) (Figure 3B). In-hospital LOS considering the units of RBC transfused during ward hospitalization also showed a significant difference between those receiving ≥3 units (18 [12; 29]) versus 1 or 2 units (10 [6; 16], p<0.01) or none (7 [4;12], p<0.01). No difference existed between patients receiving 1 or 2 units versus non-transfused (p=0.78) (Figure 3C).

## Discussion

In patients with a hemoglobin level between 70 and 90 g.L^-1^ not in life-threatening emergency situations, there was no difference in in-hospital LOS between patients transfused or not in the ED. Patients transfused in ED were more likely to experience boarding than non-ED transfused patients (OR = 2.7, CI95% [1.6; 4.5]). There was no difference in in-hospital LOS according to the number of units of RBC transfused in ED alone, yet patients receiving more units in ward and during total hospitalization had longer LOS by up to 9 days.

Few studies focus on patient outcome following transfusion in ED. One Rwandan study found that transfusing patients in ED increased mortality and in-hospital LOS [16]. However, patients included were younger, hemoglobin admission was lower and blood safety procedures differ between Europe and Africa, thus the risk of transfusion side effects might be different [17]. One study, in 12 metropolitan hospitals in France, found that acute bleeding, history of ischemic cardiopathy and older age were associated with a pre-transfusion hemoglobin level of 80 g.L^-1^, which seems consistent with our finding [12]. This study found an in-hospital LOS of 4 days. The difference of LOS could be explained by an older and sicker population and a higher ICU admission rate in our study [18, 19]. It could also be linked with a lower care access in non-metropolitan aera. ED-transfused patients were more prone to experience boarding and higher ED LOS. Even if the design of our study cannot determine whether boarding is a cause or a consequence of boarding, it is reasonable to think than transfusion in ED increase ED LOS. RBC transfusion requires a dedicated nurse for at least the first 15 minutes, and regular monitoring for 1 to 2 hours [20]. In the ED, monitoring might be impractical for nurses caring for several patients with various severity. Thus, ED-transfused patients might generate higher workloads for physicians and nurses. This heightened workload may contribute to higher in-hospital morbidity and mortality among patients with prolonged ED stays requiring hospitalization. The benefit of ED transfusion on in-hospital LOS could therefore be masked by boarding.

No protocol exists in our ED that could help to drive transfusion indications, and transfusion is at physician’s discretion. Transfusion protocols save RBC units by transfusing less RBC, improving appropriateness and developing alternative therapeutic strategies such as ferric carboxymaltose [7,21]. Other techniques to save RBC units include ED physician and nurse education about RBC transfusion [22]. One remaining challenge is to identify which patient should receive RBC transfusion in ED. Patients with history of ischemic cardiomyopathy had higher rates of transfusion in ED. However, recent studies have shown that even for patients presenting an acute coronary syndrome, a restrictive strategy did not worsen prognosis [23]. Anemia tolerance symptoms are unspecific and can be confused with other conditions. Low diastolic blood pressure could be associated with a poor tolerance of anemia [24]. Whilst our data seem to corroborate this hypothesis, with patients in the transfused group having lower diastolic blood pressure, prospective studies are needed to confirm this. New techniques, such as microcirculation evaluation could help identify patients with poor anemia tolerance. In the ICU, monitoring microcirculation before and after RBC transfusion seems promising to determine which patients could benefit the more of RBC transfusion [25].

Our data suggest that repeated transfusions in wards could be associated with a longer in-hospital LOS. In-hospital LOS was not different when considering a hemoglobin level at admission over or below 80 g.L^-1^. Dharmarajan et al. found that anemia increased LOS more in men than women [26]. We did not find differences in ED transfusion practice and in-hospital LOS regarding patient sex. A study from 2018 found that in-hospital LOS for patients with anemia was of 11 days [3]. This study found that severity of anemia was linked with in-hospital LOS, but no data concerning RBC transfusion were available. Our data showed that in patients with anemia between 70 and 90 g.L^-1^, the median LOS was 10 days, and seemed to be related with RBC transfusion during hospitalization.

We found that patients transfused in ward and receiving ≥3 RBC units in total had a longer LOS than patients transfused with ≤2 RBC units. However, the transfusion of ≥3 RBC units in ED was not associated with a significant difference of LOS. Several hypotheses could explain this observation. Firstly, patients presenting hemorrhagic shock in ED were excluded because they require immediate RBC transfusion. It is possible that some patients presented acute hemorrhage during hospitalization, which would increase LOS despite not being present at ED admission [27]. Secondly, ED physicians may initially have preferred a restrictive transfusion strategy, followed by a liberal strategy in ward [5]. Thirdly, there could be a risk of iatrogenic anemia due to multiple blood tests [28]. A vicious circle could lead to patients staying longer having more blood tests, and thus worsening hemoglobin level, requiring more RBC transfusion. A stricter control of the indications of blood tests, and lower blood volume in tests, could lower the risk of iatrogenic anemia, without impairing test quality [29].

Strengths of this study included the large total population, allowing us to conduct a propensity score on a large sample, with a good homogeneity between groups. Secondly, all consecutive patients in a calendar year were included, homogenizing the population in the standard of care. Thirdly, we included medical and surgical patients, while most studies focus on surgical patients [29]. This is of interest because ED patients fall into both fields, although medical patients are more common. The results of our study are consistent with the literature concerning LOS in anemic patients, around 11 to 14 days [3]. Finally, we excluded patients with hemorrhagic shock or needing an immediate surgical or interventional procedure, who needed an immediate transfusion regardless of their hemoglobin level.

### Limitations

Our study contains some limitations. This was a single-center study, and the local practice could probably not be extrapolated to all hospitals and areas. The retrospective setting limited the collection of some data, such as symptoms of anemia that were extracted from electronic files but could also be due to other health conditions. The hemoglobin cut-off between 70 to 90 g.L^-1^ could constitute a limit, as it is considered as severe anemia. However, recent recommendations suggest focusing more on anemia tolerance than on hemoglobin level, and that a threshold of 70 g.L-1 could be fare for most patients [6]. Hemodynamics data were not fully available, and needed to be imputed by logistical regression, which limits the validity of those data. In the excluded population, 390 (69%) were not hospitalized. A comparison of those patients with the hospitalized ones would be interesting. We did not analyze the reason for admission in both groups, assuming that it would not be linked with RBC transfusion requirement. Finally, we do not know if RBC transfusion in ED was appropriate for most patients. This is the main limitation, as the main problem of RBC transfusion in ED is transfusion appropriateness [12].

## Conclusion

In stable patients with anemia, presenting hemoglobin levels between 70 and 90 g.L^-1^, who require hospitalization, there was no benefit in terms of hospital length of stay or mortality from transfusing them in the ED rather than on the wards. However, it increased boarding. Future investigations should determine which patients should be transfused immediately in the ED to improve safety and efficiency of ED organization.

## Data Availability

All data produced in the present study are available upon reasonable request to the authors

## Acknowledgements

We thank Sarah Kabani for editing the manuscript. We thank Myriam Mezzarobba for her advices on statistical analysis.

